# Pyridostigmine in the treatment of adults with severe SARS-CoV-2 infection (PISCO): a randomised, double-blinded, phase 2/3, placebo-controlled trial

**DOI:** 10.1101/2021.04.28.21255834

**Authors:** Sergio Fragoso-Saavedra, Isaac Núñez, Belem M. Audelo-Cruz, Sarahi Arias-Martínez, Daniel Manzur-Sandoval, Alejandro Quintero-Villegas, H. Benjamín García-González, Sergio L. Carbajal-Morelos, Sergio Ponce de León-Rosales, José Gotés-Palazuelos, José A. Maza-Larrea, Yanink Caro-Vega, Isabella Batina, León Islas-Weinstein, David A. Iruegas-Nunez, Juan J. Calva, Pablo F. Belaunzarán-Zamudio, Juan Sierra-Madero, José C. Crispín, Sergio I. Valdés-Ferrer

**Author notes:** **Address correspondence to**: Sergio I. Valdés-Ferrer, Departamento de Neurología y Psiquiatría, Instituto Nacional de Ciencias Médicas y Nutrición Salvador Zubirán, Mexico City, Mexico.

## Abstract

**Background:** Hospitalised patients with severe COVID-19 have an increased risk of developing acute respiratory distress syndrome (ARDS) and death from severe systemic inflammatory response. Acetylcholine modulates the acute inflammatory response through a neuro-immune mechanism known as the inflammatory reflex. Pyridostigmine, an acetylcholine-esterase inhibitor, increases the half-life of endogenous ACh, reducing lung and systemic inflammation in murine sepsis. This trial aimed to evaluate whether pyridostigmine could decrease invasive mechanical ventilation (IMV) and death in patients with severe COVID-19.

**Methods:** We performed a parallel-group, multicentre, double-blinded, placebo-controlled, randomised clinical trial in two COVID-19-designated hospitals in Mexico City, Mexico. Adult (≥ 18-year-old), hospitalised patients with confirmed SARS-CoV-2 infection based on a positive RT-PCR test in a respiratory specimen, a computed tomography compatible with pneumonia, as well as requiring supplementary oxygen were included. Patients were randomly assigned (1:1) to receive oral pyridostigmine (60 mg per day) or placebo for a maximum of 14 days. The intention-to-treat analysis included all the patients who underwent randomisation. The primary endpoint was the composite outcome of initiation of IMV and 28-day all-cause mortality. The trial is registered in ClinicalTrials.gov, NCT04343963.

**Findings:** Between May 5, 2020, and Jan 29, 2021,188 participants were randomly assigned to placebo (n=94) or pyridostigmine (n=94). The composite outcome occurred in 22 (23·4%) *vs*. 11 (11·7%) participants, respectively (hazard ratio 0·46, 95% CI 0·22-0·96, *p*=0·03). The most frequent adverse event was diarrhoea (5 [5·3%] in the pyridostigmine group *vs* 3 [3·2%] in the placebo group). Most of the adverse events were mild to moderate, with no serious adverse events related to pyridostigmine.

**Interpretation:** Our data indicates that the addition of pyridostigmine to standard treatment reduces significantly the fatality rate among patients hospitalized for severe COVID-19.

**Funding:** Consejo Nacional de Ciencia y Tecnología, México.

## INTRODUCTION

Severe acute respiratory syndrome coronavirus 2 (SARS-CoV-2), the causative agent of coronavirus disease 2019 (COVID-19)^1^, is a novel pathogen first identified in late 2019 that reached pandemic proportions within weeks after the first cases were reported^2^. COVID-19 represents a heterogeneous disease that ranges from asymptomatic to lethal. Patients with severe COVID-19 develop a severe systemic inflammatory response^3^ and acute respiratory distress syndrome^4^, that may progress to multiple-organ failure and death^5,6^. Although the factors that determine the clinical expression of COVID-19 are not completely understood, an exuberant deregulated inflammatory response plays a central pathogenic role in patients with severe disease. In concordance, pharmacological strategies aimed at reducing inflammation have been able to moderately reduce disease outcomes. Dexamethasone, reduced mortality among patients requiring supplementary oxygen or ventilatory support. However, its use was accompanied by important and frequent adverse effects such as hyperglycaemia, psychosis, and gastrointestinal hemorrhage^7^. Tocilizumab, a recombinant humanised anti-interleukin (IL)-6 receptor monoclonal antibody, reduced the need for mechanical ventilation, as well as mortality, in patients with severe COVID-19, regardless of the degree of respiratory support needed^8^. Finally, remdesivir, an antiviral, alone or in combination with baricitinib, a selective inhibitor of Janus kinases 1 and 2, accelerated recovery and reduced in-hospital stay^9,10^. These three approaches may have increased survival rates, but severe COVID-19 remains an unsolved problem, in particular in countries with low vaccine coverage.

The central nervous system regulates the inflammatory response via the release of acetylcholine by the vagus nerve, reducing the production of inflammatory mediators, through the *inflammatory reflex*^11^. Choline-acetyltransferase (ChAT)-expressing T cells modulate inflammation via *in situ* release of acetylcholine^12^ and this effect is critical in reducing viremia in animal models of viral infection^13^. Pyridostigmine, an acetylcholinesterase inhibitor (i-ACh-e), increases acetylcholine (ACh) half-life by inhibiting its peripheral degradation. Pyridostigmine is used for the symptomatic treatment of myasthenia gravis^14^ and as pre-exposure prophylaxis for nerve gas poisoning^15^. In human immunodeficiency virus (HIV)-1 infection, pyridostigmine modulates T-cell activation and reduces circulating inflammatory markers^16–18^. Recent experimental evidence indicates that, in murine lipopolysaccharide (LPS)-induced systemic inflammation, pyridostigmine reduces lung neutrophil recruitment, as well as levels of tumour necrosis factor and IL-6 in the bronchioalveolar lavage^19^. Pyridostigmine has not yet been evaluated as immuno-modulator in acute severe human inflammatory conditions; nonetheless, indirect clinical and experimental evidence suggests that pyridostigmine may be effective in reducing the inflammation in patients with COVID-19.

Our main aim was to evaluate the efficacy of pyridostigmine as an adjunct therapy to reduce the incidence of critical illness or death in hospitalized adults with severe COVID-19.

## METHODS

### Study design and participants

We performed a double-blinded, placebo-controlled, parallel group randomised, bicentric clinical trial at Instituto Nacional de Ciencias Médicas y Nutrición Salvador Zubirán (INCMNSZ), and Instituto Nacional de Cardiología Ignacio Chávez (INCICh), two COVID-19-designated Hospitals in Mexico City, Mexico. Adult (≥18-year-old) hospitalised patients with confirmed SARS-CoV-2 infection, documented by a positive RT-PCR test for SARS-CoV-2 RNA in a respiratory specimen (nasopharyngeal or nasal swab), an imaging study compatible with pneumonia, and at least one risk factor for requiring IMV or dying (Figure 1), were invited to participate. The full list of inclusion, exclusion and elimination criteria has been published^20^.

**Figure 1.**
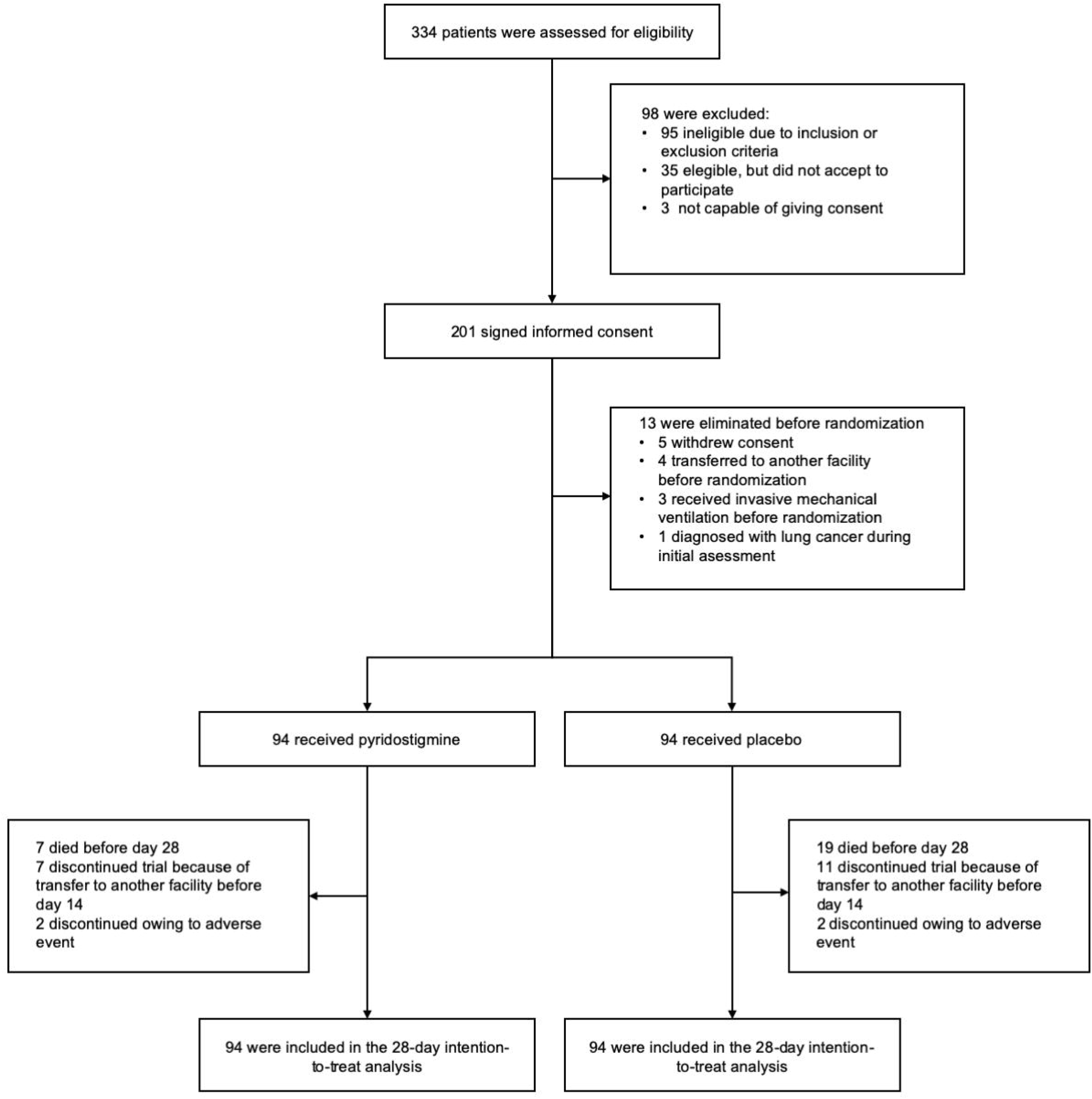
Enrolment and randomisation.

All participants provided written informed consent. The clinical study protocol was approved by the Ethics in Human Research Committees of both study centres (approval identifier: INF-3323-20-21-1).

### Randomisation and masking

Patients were randomised in a 1:1 ratio, with a parallel assignment and unstratified block-randomisation approach using a publicly available online resource (www.randomizer.org) to receive pyridostigmine or placebo besides standard medical treatment. Preparing, packing, masking, and labelling active and placebo pills, as well as overseeing drug dispensing was performed by trained pharmacy staff at both study centres. Pyridostigmine and placebo pills were unidentifiable from one another. Participants were enrolled and assigned to treatment by investigators. Throughout the study, participants, site staff and investigators were masked to the treatment allocation until completion of the intervention and follow-up phases. There was no formalised evaluation of masking.

### Procedures and outcomes

The primary outcome was a composite of invasive mechanical ventilation (IMV) or death in the 28 days following randomisation (Fig. 1), and frequency of specific adverse events. Prespecified secondary outcomes included: (a) the length of in-hospital days after randomisation; (b) hospital discharge by day-28; (c) among those who required IMV, the number of days with, and the survival rate after IMV; (d) failure to complete treatment as expected due to adverse events (other than the primary composite outcome).

Participants were allocated (1:1 ratio) to receive, in addition to standard medical treatment, either oral pyridostigmine at a dose of 60 mg/day P.O. or a matching placebo (pharmaceutical-grade starch) until the occurrence of (a) any of the prespecified outcomes, (b) hospital discharge, or (c) 14 in-hospital days. We collected demographic information from participants at baseline, including age, sex, and comorbidities (including diabetes mellitus, hypertension, obesity, cardiovascular disease, lung disease, or other chronic medical conditions). Participants already discharged from the hospital by day 28, including those who were transferred to another facility, were contacted by telephone to assess their vital and functional status.

Adverse events (AEs) were collected using data obtained from the medical health records of each study participant. The events were evaluated and coded using the Medical Dictionary and Regulatory Activities (MedDRA) version 24·0 browser and the severity of the AEs was assessed by the National Cancer Institute Common Terminology Criteria for Adverse Events, version 5·0.

Although not used throughout the study, unblinding was allowed in case of severe adverse events at the request of either the treating group of physicians or the external Data and Safety Monitoring Board (DSMB). The study was conducted in two parts, as already described^20^: a phase 2 aimed at determining safety (5 May to 4 July 2020), followed by a phase 3 aimed at evaluating the effect -or lack thereof-of pyridostigmine in patients with severe COVID-19 (5^th^ July 2020, to 30^th^ January 2021). Participants in both phases were included in the analyses of outcomes.

### Statistical analysis

The trial was planned and approved in March 2020, shortly after the first case of COVID-19 was diagnosed in Mexico. At that time, we relied on limited information about clinical outcomes in these patients. Thus, we calculated a sample size of 436 participants considering an event rate of 25% in the control group, a 10% absolute reduction (40% relative reduction) in the primary outcome as clinically significant, and a power of 80% to detect a difference in the primary outcome using a two-sided significance level of α=0·05^20^. However, we pre-stipulated that the sample size would be adjusted according to interim analyses. The first participant was recruited on May 5^th^, 2020. The DSMB performed an interim analysis after the first 44 participants (22 per group, 10% of the calculated sample) to evaluate safety on July 4^th^, 2020; a second interim analysis was conducted on December 7^th^, 2020 including 28-day outcomes on 100 participants (50 per group). Due to an observed difference in the primary outcome between groups (still blinded to investigators, but unblinded to the DSMB), as well as difficulty recruiting new participants due to a reduction in eligible patients and a swell in competing studies, we decided to stop the trial after 188 participants had been recruited (94 in each group). Considering the same α=0·05, we recalculated the statistical power of the obtained sample with the observed difference between groups (an absolute reduction of 11·7% and relative reduction of 50%), which resulted in ∼85%.

We conducted an intention-to-treat analysis for the primary outcome that included all the patients who underwent randomisation. As secondary outcomes, we assessed between-group differences of mechanical ventilation and death with or without IMV. Patients that had not developed an outcome by day 28 were censored at day 29. The magnitude of the effect of pyridostigmine on the primary outcome was estimated with a hazard ratio (HR) and its 95% confidence interval (CI) calculated with Cox proportional hazard model and the difference between groups was compared with log-rank test, as were IMV, and death with or without IMV, and discharge home although these secondary outcomes were not pre-specified in the study protocol. We built Kaplan Meier survival curves to plot the cumulative incidence of the primary outcome up to day-28. Statistical analysis was conducted with Prism GraphPad software, version 9·1·0 (GraphPad Software, San Diego, CA), and R version 4·0·0. This trial is registered in ClinicalTrials.gov, NCT04343963.

## RESULTS

### Patients

Between May 5, 2020, and Jan 29, 2021, we assessed for eligibility 334 patients, of whom 201 accepted to participate; of those, thirteen patients were excluded for the following reasons: five withdrew consent after signing but before randomisation; four were transferred to a different facility before randomisation; three required IMV before randomisation, and one was diagnosed with lung cancer (one of our exclusion criteria) during the initial assessment, therefore withdrawn from the study before randomisation. In total, 188 participants underwent randomisation, 94 in each group, all of whom received at least one dose of the assigned intervention. Enrolment and randomisation are shown in **Figure 1**. Baseline demographic and clinical characteristics were balanced between groups (**Table 1**).

**Table 1.**
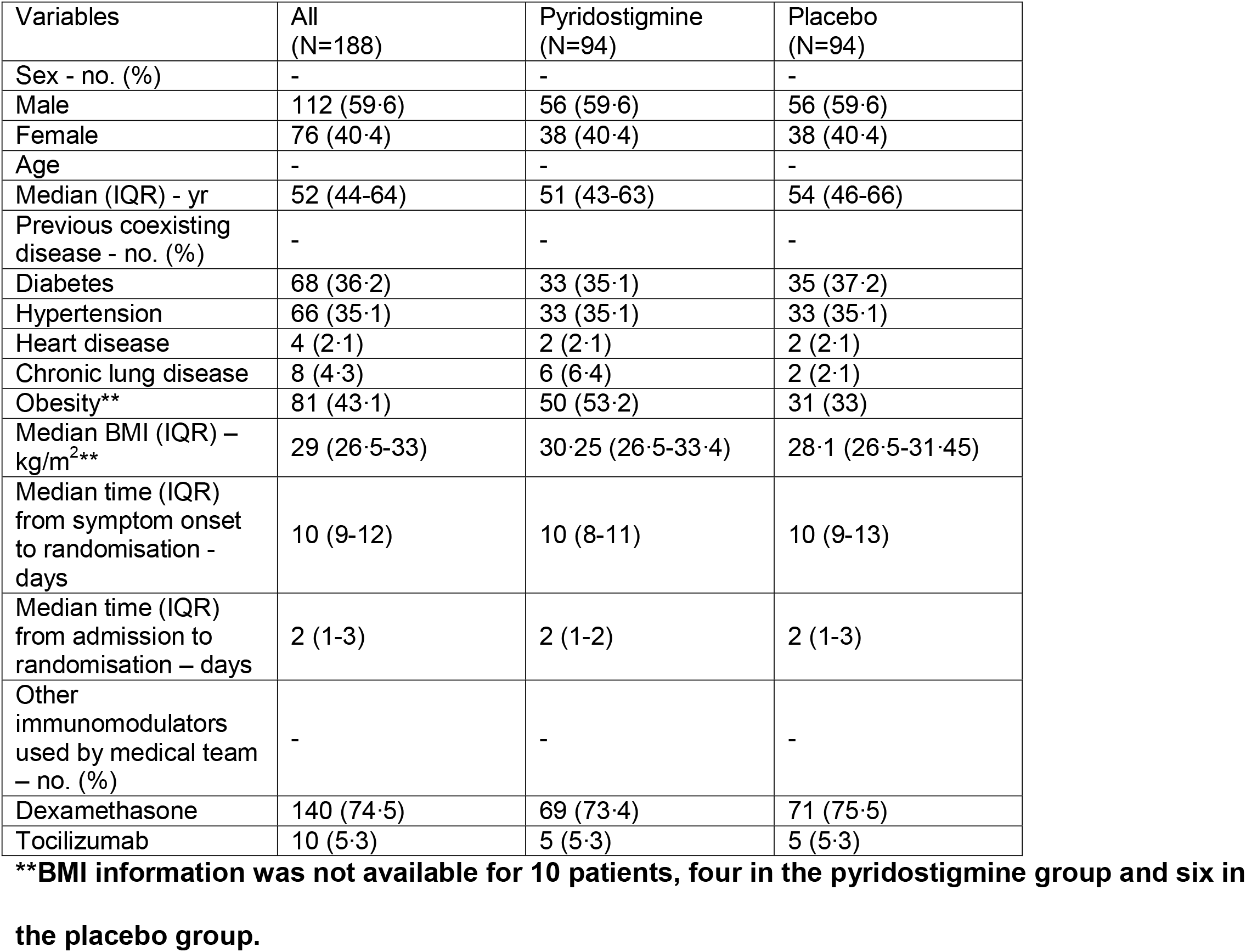
Clinical and demographical characteristics of patients at baseline.

| Variables | All<br>(N=188) | Pyridostigmine<br>(N=94) | Placebo<br>(N=94) |
| --- | --- | --- | --- |
| Sex - no. (%) | - | - | - |
| Male | 112 (59.6) | 56 (59.6) | 56 (59.6) |
| Female | 76 (40.4) | 38 (40.4) | 38 (40.4) |
| Age | - | - | - |
| Median (IQR) - yr | 52 (44-64) | 51 (43-63) | 54 (46-66) |
| Previous coexisting<br>disease - no. (%) | - | - | - |
| Diabetes | 68 (36.2) | 33 (35.1) | 35 (37.2) |
| Hypertension | 66 (35.1) | 33 (35.1) | 33 (35.1) |
| Heart disease | 4 (2.1) | 2 (2.1) | 2 (2.1) |
| Chronic lung disease | 8 (4.3) | 6 (6.4) | 2 (2.1) |
| Obesity** | 81 (43.1) | 50 (53.2) | 31 (33) |
| Median BMI (IQR) –<br>kg/m<sup>2</sup>** | 29 (26.5-33) | 30.25 (26.5-33.4) | 28.1 (26.5-31.45) |
| Median time (IQR)<br>from symptom onset<br>to randomisation -<br>days | 10 (9-12) | 10 (8-11) | 10 (9-13) |
| Median time (IQR)<br>from admission to<br>randomisation – days | 2 (1-3) | 2 (1-2) | 2 (1-3) |
| Other<br>immunomodulators<br>used by medical team<br>– no. (%) | - | - | - |
| Dexamethasone | 140 (74.5) | 69 (73.4) | 71 (75.5) |
| Tocilizumab | 10 (5.3) | 5 (5.3) | 5 (5.3) |

**\*\*BMI information was not available for 10 patients, four in the pyridostigmine group and six in** **the placebo group.**

The median age of the patients was 52 years (interquartile range [IQR], 44 to 64 years), and 59·6 % were male. The most common pre-existing conditions at enrolment were diabetes and hypertension (**Table 1**). The imbalance of obesity proportion between groups appeared to be clinically relevant (53·2% *vs*. 33%, affecting predominantly the pyridostigmine group), so we performed an *X*^*2*^ test which also showed a statistically significant difference (*p=0*·*005*). There were no relevant differences in the rest of the baseline characteristics between groups, including the number of patients who received dexamethasone or tocilizumab as part of in-hospital medical management (**Table 1**). All participants were hypoxemic and received supplementary oxygen as part of in-patient management. The median interval from symptom onset to randomisation was 10 days (IQR, 9 to 12 days). The median interval from hospital admission to randomisation was 2 days (IQR, 1 to 3 days). Dexamethasone was administered to 140 participants (69 on the pyridostigmine group and 71 on the placebo group). Tocilizumab was administered to 10 participants (5 on the pyridostigmine group, and 5 on the placebo group). None of the participants received Remdesivir.

### Primary outcome

Thirty-three participants met the primary outcome by day 28; 11 (11·7%) in the pyridostigmine group and 22 (23·4%) in the control group (HR 0·47, 95% CI 0·23-0·95, p=0·03) (**Figure 2)**. IMV was initiated in 6 (6·4%) patients in the pyridostigmine group and 7 (7·5%) in the placebo group (HR 0·81, 95%CI 0·27-2·42, p=0·7) **(Figure 3a-b)**. The median duration of IMV was 15 days (IQR, 10 to 23) in the pyridostigmine group and 17 days (IQR, 7 to 39) in the placebo group. The survival rate after IMV was 66·7% (four patients) in the pyridostigmine group and 42·9% (three patients) in the placebo group.

**Figure 2.**
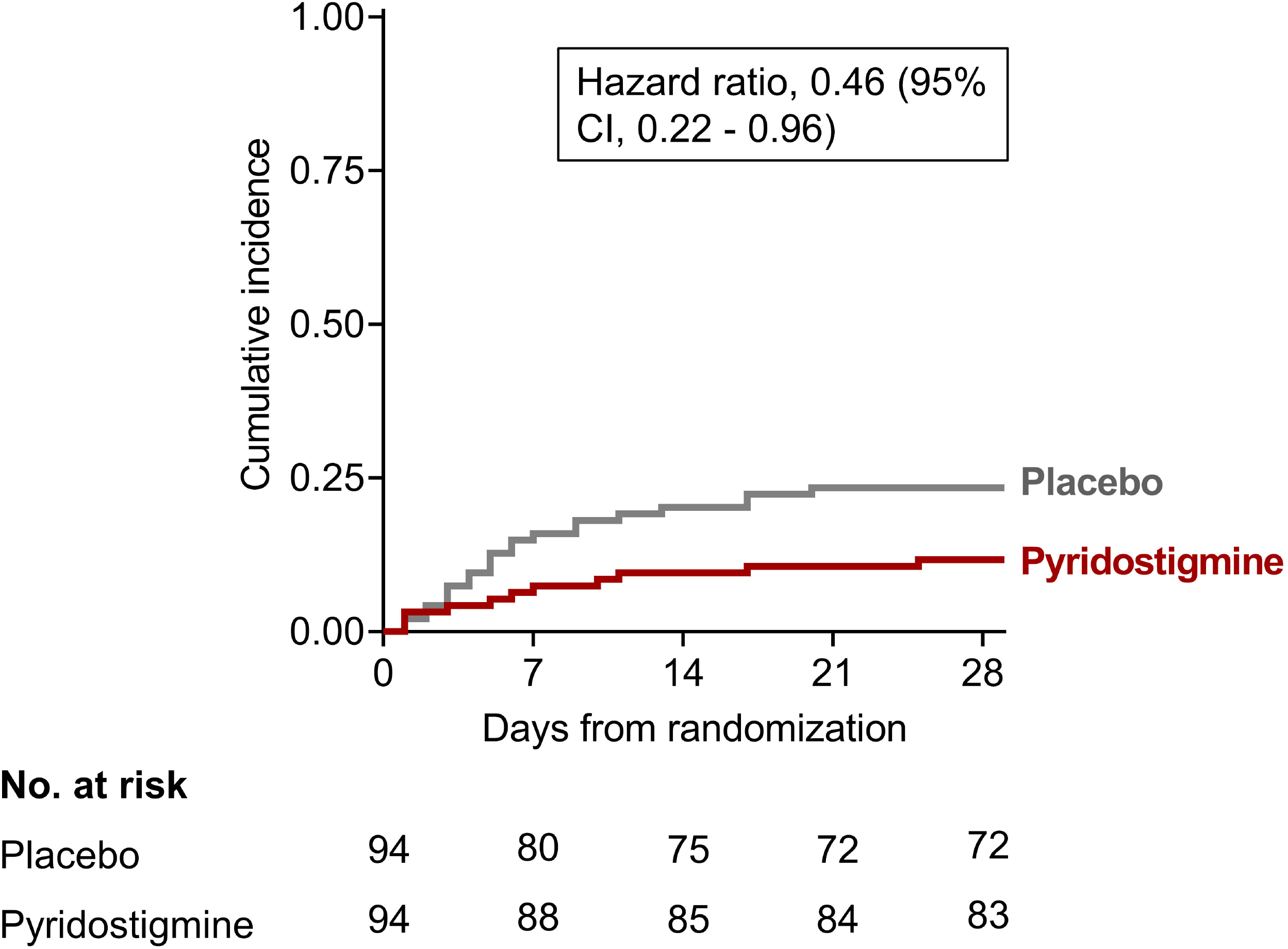
Time to Mechanical Ventilation or Death by Day 28.

**Figure 3.**
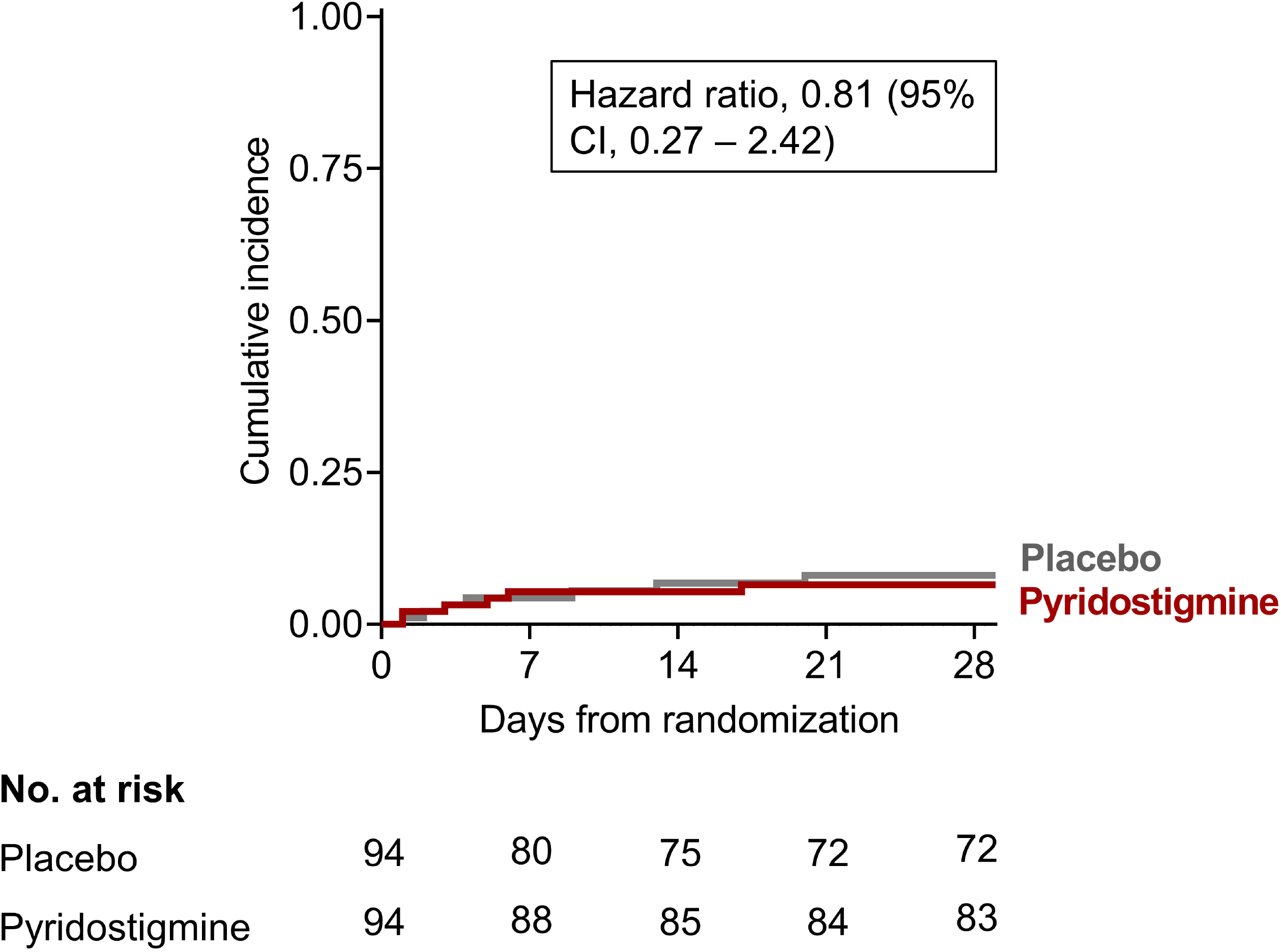
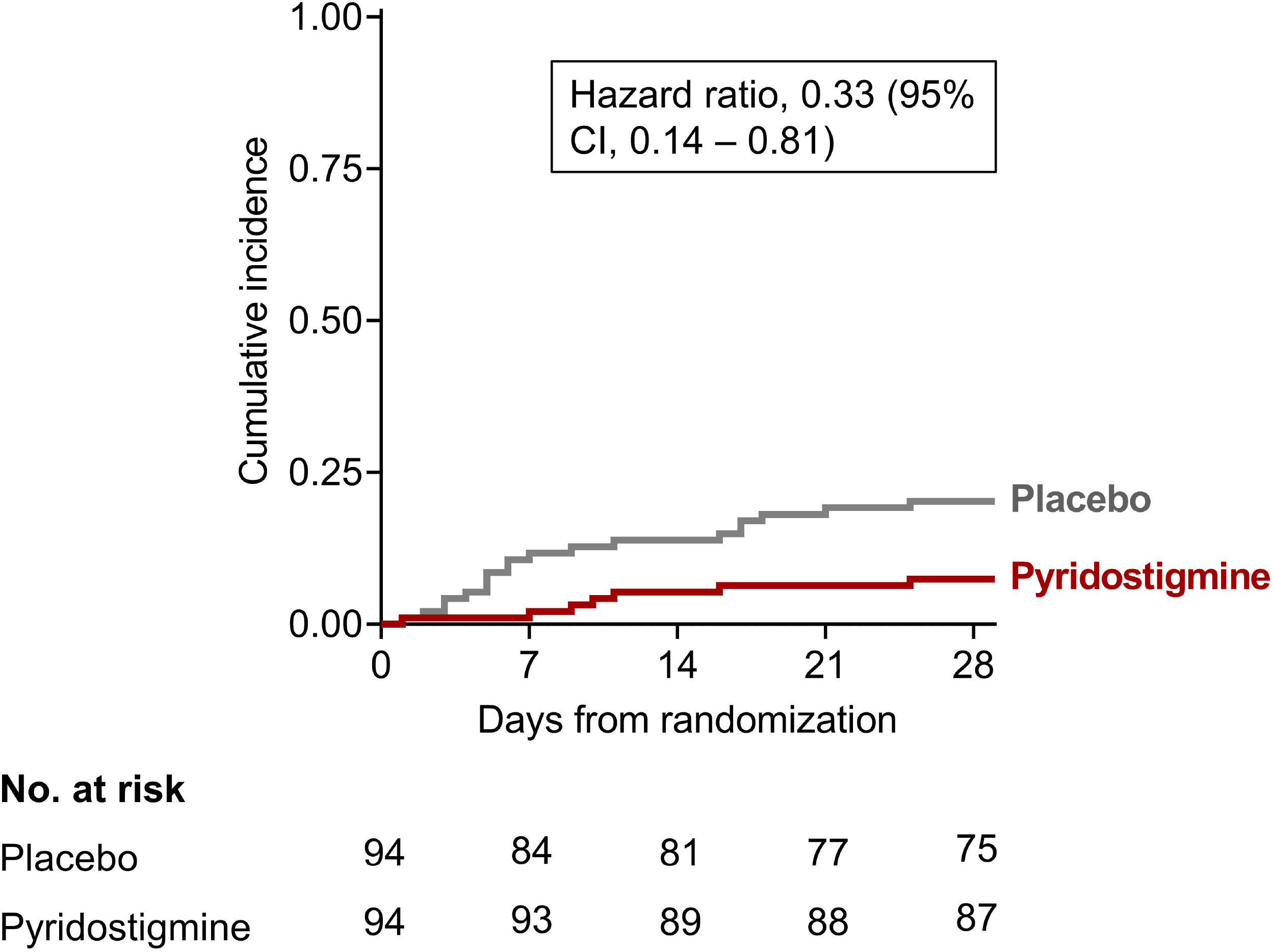
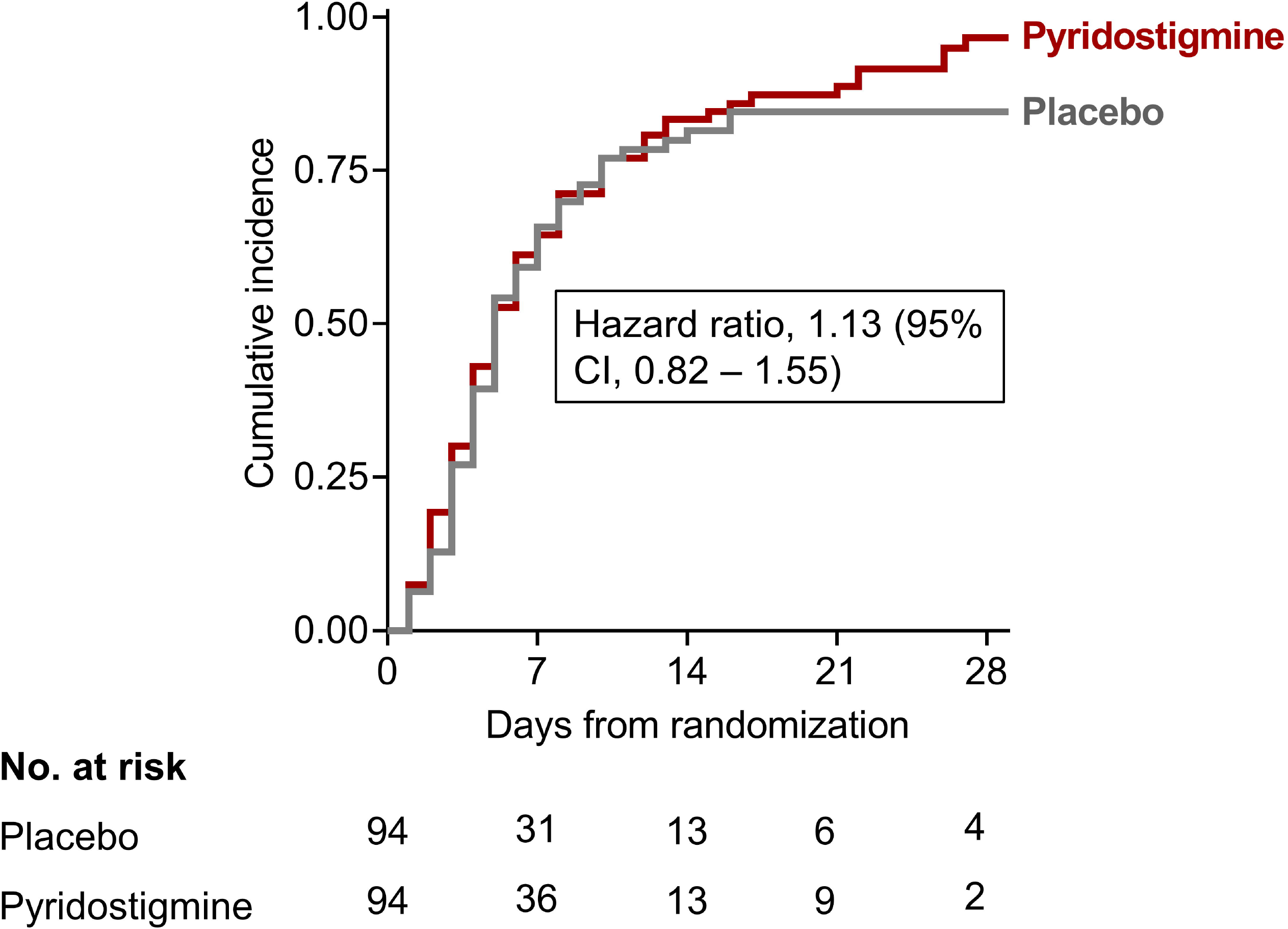
Time to Mechanical Ventilation, Death or Discharge Home by Day 28 as Separate Outcomes. A) IMV as the primary outcome; B) death as the primary outcome; and, C) Discharge home as the primary outcome. Hazard ratios were calculated with Cox proportional hazards models and p-values were obtained with a log-rank test. In the case of IMV as the primary outcome, deaths without IMV were censored. In the case of death as the primary outcome, IMV was not considered.

### Secondary outcomes

A total of 26 patients died by day 28, 7 in the pyridostigmine group and 19 in the placebo group (HR 0·33, 95%CI 0·14-0·81, p=0·01) **(Figure 3c)**. All of the deaths in the pyridostigmine group occurred in the study centre while three in the placebo group happened after the patients had been transferred to another centre. The median duration of in-hospital stay after randomisation (censored at day 29) was 5 days (IQR, 3 to 10 days) in the pyridostigmine group and 5 days (IQR, 3 to 8 days) in the placebo group. The twenty-eight-day hospital discharge rate was 90·4% (85 patients) in the pyridostigmine group and 75·5% (71 patients) in the control group (Table 2 and Fig. 3). As expected, the use of dexamethasone was adequately balanced between study groups (69 in the pyridostigmine group and 71 in the placebo group). In the placebo group, the composite primary endpoint occurred more commonly among patients that did not receive dexamethasone (35% *vs*. 20%, respectively). In contrast, in the pyridostigmine group, an opposite trend was observed (8% *vs*.13%, respectively). However, the study was not designed, nor has the power, to address drug synergism.

**Table 2.** Primary and secondary outcomes by day 28.

| Outcomes | All (N=188) | Pyridostigmine (N=94) | Placebo (N=94) | HR (95% CI) | P-value |
| --- | --- | --- | --- | --- | --- |
| Primary outcome |  |  |  |  |  |
| Invasive mechanical ventilation or death – no. (%) | 33 (17.5%) | 11 (11.7%) | 22 (23.4%) | 0.46 (0.23-0.95) | 0.03 |
| Secondary outcomes |  |  |  |  |  |
| Invasive mechanical ventilation – no. (%) | 13 (6.9%) | 6 (6.3%) | 7 (7.4%) | 0.81 (0.27-2.42) | 0.7 |
| Median time (IQR) in ICU with invasive mechanical ventilation - days | 17 (11-23) | 15 (15-21) | 17 (12-30) | - | - |
| Survival after IMV – no. (%) | 7 (53.8) | 4 (66%) | 3 (42.9) | - | - |
| Death - no. (%) | 26 (13.8%) | 7 (7.4%) | 19 (20.2%) | 0.33 (0.14-0.81) | 0.01 |
| *Median time (IQR) from randomisation to discharge home – days | 5 (3-9) | 5 (3-10) | 5 (3-8) | - | - |
| Discharge home by day 28 – no. (%) | 156 (82.9) | 85 (90.4) | 71 (75.5) | 1.13 (0.82-1.55) | 0.4 |
| Adverse events - no. (%) | 83 (44.1) | 45 (47.9) | 38 (40.4) | - |  |
| Failure to complete treatment due to adverse effects – no. (%) | 4 (2) | 2 (2) | 2 (2) | - | - |

\*For time to discharge home in patients that were transferred, the time from randomisation until discharge home from the hospital that they were transferred to was used. Two patients did not have this information available but were discharged home alive. For them, the date of transfer was used. Patients that were still in the hospital on day 28 were censored on day 29.

We collected and analysed 83 AEs throughout the study. Overall, 45 (47·9%) patients of the pyridostigmine group and 38 (40·4%) of the placebo group had at least one adverse drug reaction. The rate and severity of AEs were similar between both groups (Supplemental Tables 1-3).

One hundred and sixty-six (88·3%) participants received experimental treatment (i.e., pyridostigmine or placebo) as assigned during their in-hospital stay; 22 (11·7%) discontinued treatment early, nine (9·5%) in the pyridostigmine group, and 13 (13·8%) in the placebo group. Two participants in each group discontinued trial medication due to an adverse event other than the principal outcome: one patient presented nausea and one mild abdominal pain in the pyridostigmine group (known adverse effects of pyridostigmine), and in the placebo group one patient had pyrosis and one presented diarrhoea. The remaining patients discontinued trial medication after being transferred to another facility **(Figure 1)**. Drug adherence was supervised by a physician or nurse, and each administration was confirmed in the electronic records; therefore, among the 166 participants who received medication as assigned, adherence was 100%.

Finally, as obesity status differed between study groups, we performed a sensitivity analysis with a Cox proportional hazards model adjusted for obesity status. We excluded 10 patients (four from the pyridostigmine group and six from the control group) that had no information on obesity status (of which only one had a primary outcome) for this sensitivity analysis, obtaining an adjusted HR of 0·42 (95% CI 0·19-0·91, p=0·05) for the primary outcome, 0·78 (95%CI 0·24-2·54, p=0·3) for IMV, and 0·32 (95%CI 0·13-0·77, p=0·03) for death.

## DISCUSSION

Here, we show that added to standard care, pyridostigmine is associated with decreased IMV and mortality in patients hospitalized for severe COVID-19. This was driven by a decrease in all-cause mortality in the pyridostigmine group compared to that of the placebo group (by 12·7 percentage points). IMV was an infrequent outcome, which explains the lack of difference when analysing this outcome by itself. Also, the adjunct use of pyridostigmine increased the probability of being discharged from the hospital alive within 28 days.

Early reports estimated mortality rates as elevated as 49% for hospitalized patients with severe COVID-19^21^. Although the in-hospital mortality rate has been declining throughout the pandemic, recent studies still show rates ranging from 20 to over 40%^22–25^. For instance, the risk of death still exceeded 20% in the dexamethasone and tocilizumab arms of the RECOVERY trial, being the only two treatments that have shown a reduction in mortality^7,24^. Similarly, in the placebo group we observed a 28-day mortality of 20.21%, but a majority had received dexamethasone as part of the standard of care, indicating that, without further intervention, in-hospital mortality of severe COVID-19 remains high. The severity and mortality of COVID-19 are mediated by the development of an uncontrolled inflammatory response to infection; elevated levels of proinflammatory cytokines and other immune mediators have been associated with multiorgan failure due to endothelial damage and tissue injury^4,6^. Hence, finding novel immunomodulatory strategies represents a promising strategy to reduce the severity and mortality of COVID-19. That, along with the repurposing of drugs with well-characterized safety profiles and readily available production lines, might lead to faster development of anti-COVID-19 therapies if proven efficacious in well-designed, randomised clinical trials.

In mammals, the central nervous system has mechanisms to control the inflammatory response. During inflammatory states, the vagus nerve can inhibit the synthesis and release of inflammatory cytokines, thereby reducing both local damage and mortality secondary to severe systemic inflammation in murine models as diverse as sepsis, ischemia, and reperfusion damage, or obesity^26–29^. The vagus nerve can be stimulated electrically and chemically. Chemical stimulation using cholinergic agonists has shown promising effects in murine and cellular models of inflammation^29,30^. Acetylcholine esterase inhibitors (i-ACh-e) are a family of drugs used regularly by millions of patients, including older adults with Alzheimer’s disease and other dementias, as well as patients with myasthenia gravis and dysautonomia^14,31–34^. These drugs inhibit the enzymatic degradation of endogenous ACh, resulting in greater bioavailability and, therefore, increasing the possibility of binding to both nicotinic and muscarinic receptors. In addition to the approved uses of i-ACh-e in human pathology, there is evidence in various murine models of their efficacy in experimental sepsis and severe inflammatory response^19,27,28,30^, suggesting that i-ACh-e drugs may exert a potential immunomodulatory effect in patients with severe systemic inflammatory response syndrome.

Regarding safety concerns, at the used dose of pyridostigmine (60 mg/day), the rate of adverse events we observed was 47.9% -none severe- and not different from the observed rate in the placebo group (40.4%).

The reduction in mortality in patients with severe COVID-19 who received pyridostigmine may be due to an immunomodulatory effect, however, its precise biological effect in this novel disease remains to be studied. Different immunomodulatory strategies have been tested that have sought to selectively inhibit the biological activity of certain pro-inflammatory cytokines, such as IL-6^35^; however, the global inhibition of several of them by pharmacologically stimulating the inflammatory reflex could be more effective, which is the effect sought with the administration of pyridostigmine. We are currently working in understanding the mechanism behind our observed effect.

Our study has important limitations. The originally planned sample size could not be achieved. Still, a significant difference was observed because of a greater than expected effect. Also, some patients requiring IMV could probably not receive it because of either, a pre-existing living will; last-minute patient (or proxy) refusal of intubation; or, intermittently throughout the study period, lack of availability of critical care beds^22,23^. Our primary outcome included initiation, but not necessarily the requirement, of IMV. This is reflected by the fact that the bulk of the primary outcome was due to deaths among participants who did not receive IMV, most likely related to lack of critical care space. Thus, our results may not be generalizable to settings that do not have this problem. While most baseline characteristics were evenly balanced between groups, obesity (a known predictor of adverse outcomes in COVID-19) was randomly more prevalent in the pyridostigmine group; this imbalance that in principle plays against pyridostigmine strengthens our observed outcomes. As a proof-of-concept, this study needs to be replicated or refuted in independent clinical trials. Also, at this point, our results cannot be extrapolated to patients with less severe disease, or those already receiving IMV, without prior verification in clinical trials. Finally, medium-term outcomes (90 days) will be reported at a later time point.

The strengths of the study include that it is a double-blinded, placebo-controlled, multicentric randomised clinical trial that evaluated a therapeutic intervention that is inexpensive, has a positive pharmacological profile, is safe, and is widely available as a generic drug. The demographics of our study population, including age and comorbid conditions, are representative of a large group of the underserved population worldwide (particularly across the Americas), a population in urgent need of safe, effective, life-saving, and affordable treatments for severe COVID-19.

In conclusion, we present evidence to support the use of pyridostigmine as an add-on treatment to standard medical care in patients hospitalized for severe COVID-19. Our data indicate that this inexpensive and readily available drug may significantly reduce mortality without imposing relevant adverse effects.

## Data Availability

Deidentified data will be made available upon request to the corresponding author

## Contributors

JCC, PFBZ, JSM and SIVF designed the study. SFS, SIVF, and IN wrote the manuscript. JCC, JSM and JJC contributed to the writing of the article. SFS did the block randomisation. SFS, BMAC, SAM, DMS, IN, HBGG, AQV, SLCM and SIVF contributed to the recruitment of participants. JAML and JGP evaluated the safety of the pharmacological intervention. SFS, BMAC, DMS, DAIN, IB, and LIW contributed to data gathering and entry. SFS, IN, SIVF analysed all data. PFBZ, YCV, and SPdeLR provided additional statistical support. SFS, IN, and SIVF accessed and verified the trial data. All authors approved the submission of this article.

## Data sharing

De-identified individual participant data and all protocol documents can be provided to others upon reasonable request on approval of a written request to the corresponding author.

## Acknowledgements

This study was funded by Consejo Nacional de Ciencia y Tecnología, Mexico (COVID-19 Fund [F0005-2020-01]; grant 311790), to SIV-F]. The authors are grateful to Kevin J. Tracey, M.D., for his insight and suggestions that positively impacted this manuscript; to Claudia Quiñones, B.Ch. and, Elia Criollo-Mora, B.Ch. for preparing, packing, masking, labelling, and dispensing treatments; to Dr Virginia Pascual-Ramos for providing ethical and statistical recommendations throughout the study; and to Dr Florencia Rosetti and the Immunopathology Lab, INCMNSZ for their logistic support. Lastly, the authors wish to dedicate this manuscript to all medical and surgical residents and fellows across the world for their generous and selfless work during the COVID-19 pandemic. We understand that this is not the kind of residency training you dreamed of, but your work has, time and again, made a difference and every day you make us all proud.

## References

1 Zhu N, Zhang D, Wang W, et al. A Novel Coronavirus from Patients with Pneumonia in China, 2019. N Engl J Med 2020; 382: 727–33.

2 Johns Hopkins Coronavirus Resource Center. https://coronavirus.jhu.edu/map.html (accessed May 15, 2021).

3 Xu Z, Shi L, Wang Y, et al. Pathological findings of COVID-19 associated with acute respiratory distress syndrome. Lancet Respir Med 2020; 8: 420–2.

4 Huang C, Wang Y, Li X, et al. Clinical features of patients infected with 2019 novel coronavirus in Wuhan, China. Lancet 2020; 395: 497–506.

5 Ruan Q, Yang K, Wang W, Jiang L, Song J. Clinical predictors of mortality due to COVID-19 based on an analysis of data of 150 patients from Wuhan, China. Intensive Care Med 2020; 46: 846–8.

6 Mehta P, McAuley DF, Brown M, Sanchez E, Tattersall RS, Manson JJ. COVID-19: consider cytokine storm syndromes and immunosuppression. Lancet 2020; 395: 1033–4.

7 Horby P, Lim WS, Emberson J, et al. Dexamethasone in Hospitalized Patients with Covid-19. N Engl J Med 2021; 384: 693–704.

8 RECOVERY Collaborative Group. Tocilizumab in patients admitted to hospital with COVID-19 (RECOVERY): preliminary results of a randomised, controlled, open-label, platform trial. Lancet 2021; 397: 1637–45.

9 Beigel JH, Tomashek KM, Dodd LE, et al. Remdesivir for the Treatment of Covid-19 — Final Report. N Engl J Med 2020; 383: 1813–26.

10 Kalil AC, Patterson TF, Mehta AK, et al. Baricitinib plus Remdesivir for Hospitalized Adults with Covid-19. N Engl J Med 2021; 384: 795–807.

11 Borovikova L V., Ivanova S, Zhang M, et al. Vagus nerve stimulation attenuates the systemic inflammatory response to endotoxin. Nature 2000; 405: 458–62.

12 Rosas-Ballina M, Olofsson PS, Ochani M, et al. Acetylcholine-synthesizing T cells relay neural signals in a vagus nerve circuit. Science (80-) 2011; 334. DOI:10.1126/science.1209985.

13 Cox MA, Duncan GS, Lin GHY, et al. Choline acetyltransferase–expressing T cells are required to control chronic viral infection. Science (80-) 2019; 363: 639–44.

14 Gilhus NE, Verschuuren JJ. Myasthenia gravis: Subgroup classification and therapeutic strategies. Lancet Neurol 2015; 14: 1023–36.

15 Keeler JR, Hurst CG, Dunn MA. Pyridostigmine Used as a Nerve Agent Pretreatment Under Wartime Conditions. JAMA J Am Med Assoc 1991; 266: 693–5.

16 Valdés-Ferrer SI, Crispín JC, Belaunzarán PF, Cantú-Brito CG, Sierra-Madero J, Alcocer-Varela J. Acetylcholine-esterase inhibitor pyridostigmine decreases T cell overactivation in patients infected by HIV. AIDS Res Hum Retroviruses 2009; 25. DOI:10.1089/aid.2008.0257.

17 Valdés-Ferrer SI, Crispín JC, Belaunzarán-Zamudio PF, et al. Add-on Pyridostigmine enhances CD4+ T-cell recovery in HIV-1-infected immunological non-responders: A proof-of-concept study. Front Immunol 2017; 8: 1–6.

18 Robinson-Papp J, Nmashie A, Pedowitz E, et al. The effect of pyridostigmine on small intestinal bacterial overgrowth (SIBO) and plasma inflammatory biomarkers in HIV-associated autonomic neuropathies. J Neurovirol 2019; 25: 551–9.

19 Bricher Choque PN, Vieira RP, Ulloa L, et al. The Cholinergic Drug Pyridostigmine Alleviates Inflammation During LPS-Induced Acute Respiratory Distress Syndrome. Front. Pharmacol.. 2021; 12: 966.

20 Fragoso-Saavedra S, Iruegas-Nunez DA, Quintero-Villegas A, et al. A parallel-group, multicenter randomized, double-blinded, placebo-controlled, phase 2/3, clinical trial to test the efficacy of pyridostigmine bromide at low doses to reduce mortality or invasive mechanical ventilation in adults with severe SARS-CoV-2 inf. BMC Infect Dis 2020; 20: 765.

21 Wu Z, McGoogan J. Characteristics of and important lessons from the coronavirus disease 2019(COVID-19) outbreak in China. Jama 2020; 2019: 10.1001/jama.2020.2648.

22 Rossman H, Meir T, Somer J, et al. Hospital load and increased COVID-19 reñated mortality in Israel. Nat Commun 2021; 12: 1–7.

23 Olivas-Martínez A, Cárdenas-Fragoso JL, Jiménez JV, et al. In-hospital mortality from severe COVID-19 in a tertiary care center in Mexico City; causes of death, risk factors and the impact of hospital saturation. PLoS One 2021; 16: e0245772.

24 de Souza FSH, Hojo-Souza NS, Batista BD de O, da Silva CM, Guidoni DL. On the analysis of mortality risk factors for hospitalized COVID-19 patients: A data-driven study using the major Brazilian database. PLoS One 2021; 16: e0248580.

25 Núñez I, Priego-Ranero ÁA, García-González HB, et al. Common hematological values predict unfavorable outcomes in hospitalized COVID-19 patients. Clin Immunol 2021; 225: 108682.

26 Yeboah MM, Xue X, Duan B, et al. Cholinergic agonists attenuate renal ischemia-reperfusion injury in rats. Kidney Int 2008; 74: 62–9.

27 Satapathy SK, Ochani M, Dancho M, et al. Galantamine alleviates inflammation and other obesity-associated complications in high-fat diet-fed mice. Mol Med 2011; 17. DOI:10.2119/molmed.2011.00083.

28 Zaghloul N, Addorisio ME, Silverman HA, et al. Forebrain cholinergic dysfunction and systemic and brain inflammation in murine sepsis survivors. Front Immunol 2017; 8. DOI:10.3389/fimmu.2017.01673.

29 Lehner KR, Silverman HA, Addorisio ME, et al. Forebrain cholinergic signaling regulates innate immune responses and inflammation. Front Immunol 2019; 10: 1–11.

30 Rosas-Ballina M, Valdés-Ferrer SI, Dancho ME, et al. Xanomeline suppresses excessive pro-inflammatory cytokine responses through neural signal-mediated pathways and improves survival in lethal inflammation. Brain Behav Immun 2015; 44: 19–27.

31 Mayeux R. Clinical practice. Early Alzheimer’s disease. N Engl J Med 2010; 362: 2194–201.

32 Querfurth HW, Laferla FM. Alzheimer’s Disease. N Engl J Med 2018; 326: 329–44.

33 O’Brien JT, Thomas A. Vascular dementia. Lancet 2015; 386: 1698–706.

34 Singer W, Sandroni P, Opfer-Gehrking TL, et al. Pyridostigmine treatment trial in neurogenic orthostatic hypotension. Arch Neurol 2006; 63: 513–8.

35 Salama C, Han J, Yau L, et al. Tocilizumab in Patients Hospitalized with Covid-19 Pneumonia. N Engl J Med 2021; 384: 20–30.

