## Supplemental material for "Pyridostigmine in the treatment of adults with severe SARS-CoV-2 infection (PISCO): a randomised, double-blinded, phase 2/3, placebo-controlled trial" (from another version of this preprint: version 1, 2021-04-29)

| Variable | Pyridostigmine (n=94)  No. (%) | Placebo (n=94)  No. (%) | Total (n=188) |
| --- | --- | --- | --- |
| Total AEs | 45 (47.9) | 38 (40.4) | 83 (44.1) |
| Participants with one AE* | 36 (38.3) | 31 (33) | 67 (35.6) |
| Participants with 2 or more AEs* | 9 (9.6) | 7 (7.4) | 16 (8.51) |
| Participants with at least one serious AE* | 3 (3.2) | 3 (3.2) | 6 (3.19) |
| Participants with at least one AE related to the treatment* | 18 (19.4) | 11 (11.7) | 29 (30.8) |
| Participants with at least one serious AE related to the treatment* | 0 | 0 | 0 |
| Grade 1 and 2 AEs | 42 (93.3) | 35 (92.1) | 77 (92.7)‡ |
| Grade 3 and 4 AEs | 3 (6.6) | 3 (7.89) | 6 (7.22)‡ |
| **Supplementary table 1. Summary of the adverse events in the study population.** AE: Adverse event. *Participants are counted once for each category. ‡Proportion from the total number of adverse events. | | | |

| System Organ Class | Preferred Term | Pyridostigmine (n=94)  No. (%) | Placebo (n=94)  No. (%) |
| --- | --- | --- | --- |
| Cardiac disorders | Acute myocardial infarction | - | 1 (1) |
|  | Supraventricular tachycardia | - | 1 (1) |
| Injury, poisoning and procedural complications | Inadequate hemodialysis | 1 | 1 (1) |
| Renal and urinary disorders | Acute kidney injury | 1 (1) | - |
| Respiratory, thoracic and mediastinal disorders | Pulmonary embolism | 1 (1) | - |
| **Supplementary table 2.** Description of serious adverse events according to MEdDRA by treatment group | | | |

| System Organ Class | Preferred Term | Pyridostigmine (n=94)  No. (%) | Placebo (n=94)  No. (%) |
| --- | --- | --- | --- |
| Blood and lymphatic system disorders | Leukocytosis | - | 4 |
| Cardiac disorders | Bundle branch block right | 1 | - |
|  | Sinus bradycardia | 3 | 4 |
|  | Tachycardia | 1 | - |
| Gastrointestinal disorders | Abdominal distension | 1 | - |
|  | Abdominal pain | - | 1 |
|  | Abdominal pain upper | 1 | - |
|  | Aphthous ulcer | - | 1 |
|  | Constipation | - | 1 |
|  | Diarrhoea | 5 | 3 |
|  | Dyspepsia | 2 | - |
|  | Nausea | 1 | 1 |
|  | Salivary hypersecretion | 1 | - |
|  | Vomiting | 1 | - |
| General disorders and administration site conditions | Chest pain | 2 | - |
|  | Fatigue | 1 | - |
| Injury, poisoning and procedural complications | Fall | 1 | - |
| Investigations | Transaminases increased | 1 | 3 |
| Metabolism and nutrition disorders | Hyperglycemia | 7 | 1 |
|  | Hypoglycemia | - | 1 |
|  | Hyponatremia | - | 4 |
|  | Decreased appetite | 1 | - |
| Nervous system disorders | Somnolence | - | 2 |
| Psychiatric disorders | Anxiety | 3 | 1 |
|  | Delirium | 4 | 2 |
| Renal and urinary disorders | Acute kidney injury | - | 2 |
| Skin and subcutaneous tissue disorders | Angioedema | 1 | - |
|  | Hyperhidrosis | 1 | - |
|  | Urticaria | 1 | - |
| Vascular disorders | Hypertension | 1 | 3 |
|  | Hypotension | 1 | 1 |
| Total |  | 42 | 35 |
| **Supplementary table 3.** Description of non-serious adverse events according to MEdDRA by treatment group. No, Number of subjects with at least one adverse event. | | | |

**Definition of adverse event (AE)**

Any untoward medical occurrence in the study participant, that is considered to be temporally associated with the use of the study treatment, whether or not is deemed to be related to the study treatment. An adverse event can be any unfavorable and unintended abnormal laboratory finding, symptom, new or exacerbated or disease temporally associated with the use of the study treatment.

The definition of AE does not consider an event related to the disease being studied or expected progression, signs, or symptoms of this disease, unless it´s more severe than expected for the participant’s condition.

**Definition of serious adverse event (SAE)**

Any untoward medical occurrence in the study participant that results in:

- Death
- Is considered to be life-threatening (the condition of the study participant was considered to be at risk at the moment of the event).
- Generates inpatient hospitalization or prolongation of the existing hospitalization.
- Results in permanent disability or permanent organ damage.
- Results in a congenital defect or abnormality.
- Other situations where the investigators or the attending physician consider that an urgent medical intervention is required in order to avoid the appearance of any of the former events.
